# SARS-CoV-2 RT-PCR profile in 298 Indian COVID-19 patients : a retrospective observational study

**DOI:** 10.1101/2020.06.19.20135905

**Authors:** Bisakh Bhattacharya, Rohit Kumar, Ved Prakash Meena, Manish Soneja, Saurabh Vig, Vandana Rastogi, Sushma Bhatnagar, Anant Mohan, Naveet Wig

## Abstract

**Background:** Despite being in the 5th month of pandemic, knowledge with respect to viral dynamics, infectivity and RT-PCR positivity continues to evolve.

**Aim:** To analyse the SARS CoV-2 nucleic acid RT-PCR profiles in COVID-19 patients.

**Design:** It was a retrospective, observational study conducted at COVID facilities under AIIMS, New Delhi.

**Methods:** Patients admitted with laboratory confirmed COVID-19 were eligible for enrolment. Patients with incomplete details, or only single PCR tests were excluded. Data regarding demographic details, comorbidities, treatment received and results of SARS-CoV-2 RT-PCR performed on nasopharyngeal and oropharyngeal swabs, collected at different time points, was retrieved from the hospital records.

**Results:** 298 patients were included, majority were males (75·8%) with mean age of 39·07 years (0·6-88 years). The mean duration from symptom onset to first positive RT-PCR was 4·7 days (SD 3·67), while that of symptom onset to last positive test was 17·83 days (SD 6·22). Proportions of positive RT-PCR tests were 100%, 49%, 24%, 8·7% and 20·6% in the 1st, 2nd, 3rd, 4^th^ & >4 weeks of illness. 12 symptomatic patients had prolonged positive test results even after 3 weeks of symptom onset. Age >= 60 years was associated with prolonged RT-PCR positivity (statistically significant).

**Conclusion:** This study showed that the average period of PCR positivity is more than 2 weeks in COVID-19 patients; elderly patients have prolonged duration of RT-PCR positivity and requires further follow up.

## Introduction

The COVID-19 pandemic continues to affect millions of people worldwide with more than 50,000 active cases in India and around 3000 deaths. ^(1)^ Identifying potentially infectious patients and isolating them is critical in order to halt the spread of infection. As per WHO guidance all suspected cases should be screened by nucleic acid amplification tests (NAATs) such as the reverse-transcription polymerase chain reaction (RT-PCR) on upper and lower respiratory tract samples.^(2)^ Serological assays can aid in the diagnosis of suspected COVID-19 cases but is not available as yet and routine viral culture is not recommended. Hence currently the RT-PCR assay forms the backbone of SARS-CoV-2 diagnosis. However the dynamics of viral shedding has not been fully understood. Currently as per the revised government of India guidelines asymptomatic or mildly symptomatic patients can be discharged 10 days after symptom onset and the strategy of repeat RT-PCR testing has been done away with.^(3)^ In this retrospective study we aim to characterise the RT-PCR profiles of Indian COVID-19 patients which can further aid in defining an appropriate time for discharge or isolation of infected patients.

## Methods

### Study design & setting

This study was conducted at AIIMS, New Delhi, a tertiary care teaching hospital with special dedicated wards and isolation centers for COVID-19 patients. It was an observational retrospective study where all patients who were admitted with a diagnosis of COVID-19 according to the government of India guidelines between 20th March 2020 to 30th April 2020 were included.

### Procedure & data collection

All patients admitted, between 20th March 2020 to 30th April 2020 at AIIMS, New Delhi were screened for eligibility. Cases with laboratory confirmed diagnosis of COVID-19, made by positive SARS-CoV-2 RT-PCR on nasopharyngeal samples, were enrolled regardless of symptomatology. Patients with at least 2 SARS-CoV-2 RT-PCR reports were included. Patient with incomplete case records; unavailable test results or only single tests were excluded. Details including demographic details, comorbidities, clinical features (symptomatology, date of onset of illness), date of Ist and subsequent PCR tests) were noted retrospectively from hospital records. For symptomatic patients, duration will be considered from day of symptom onset, while for asymptomatic patients, duration from first RT-PCR will be considered. Patients were eligible for discharge after they became asymptomatic and had two reports with RT -PCR results as negative (tests performed at least 24 hours apart).

### Statistical analysis

The statistical software STATA v15.1 (STATA corp, LLC, TX, USA) was used for all statistical analysis. For testing of hypothesis two tailed test was considered and p value of less than 0.05 was considered to be statistically significant. Descriptive analysis of the data was carried out first to summarise the study population. Mean ± Standard Deviation were given for continuous variables with no extreme values. Continuous variables with extreme value(skewed) were given by Median and IQR and qualitative variables were expressed as proportion (%). Data normality is checked by Normal probability plot and Kolmogorov - Smirnov test. For comparison of two groups Student’s t test was performed and for the comparison of more than two groups one way ANOVA followed by Bonferroni correction for multiple comparisons was applied.

## Results

324 patients were eligible for enrolment, and 26 cases were excluded (Figure 1). A total of 298 patients were included in the study for final analysis. 164 (55%) patients were symptomatic and 134 (45%) were asymptomatic; most symptomatic patients having mild or moderate disease as per ICMR (Indian Council of Medical Research) guidelines. ^(4)^ All the patients were managed as per ICMR guidelines and institution protocol. Three patients had severe illness and all of them succumbed to illness (they were excluded from the study as only 1 RT-PCR result was available). The mean age of our study population was 39·07 years (6 months-88 years); majority cases were males 75·8% (226) males. 39 (13·08%) patients had diabetes mellitus and 23 (7·7%) had hypertension.

**Figure 1.**
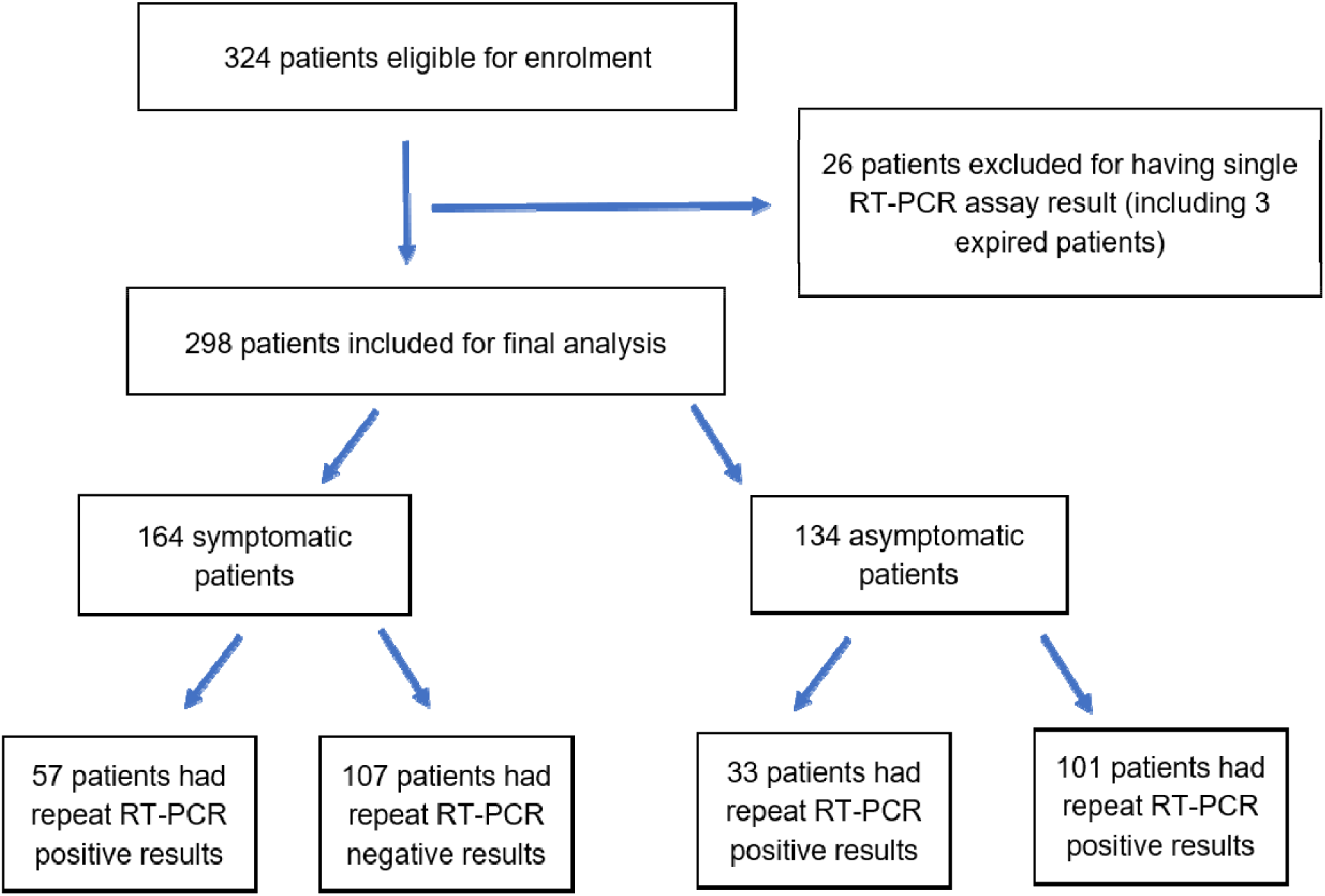
Flow diagram of the study

A total of 930 RT-PCR tests were done for 298 patients, with an average of 3·12 tests per patient. 252 patients (137 symptomatic, 115 asymptomatic) had been declared as cured at the time of final analysis. A total of 90 patients had a second positive RT-PCR assay whereas the rest 208 had either repeat negative assays or repeat test results were unavailable for them. The mean duration to first positive RT-PCR from onset of symptoms was 4·7 days (1-21 days; SD 3·67); the median duration was 3 days (IQR:2-7). The mean duration from symptom onset to last positive RT-PCR was 17·83 (8-33 days; SD 6·22), median duration was 18 days (IQR 13·75-21). The percentage of positive RT-PCR tests in symptomatic patients was highest in the 1st week (100%), followed by 49%, 24%, 8·7% and 20·6% in the 2nd, 3rd, 4th week and beyond 4th week respectively (Figure 2). The percentage of positive RT-PCR results following 1st RT-PCR test (regardless of symptoms) was 50% in 1st week, followed by 28%, 10·8%, 16·2% and 6.6% in 2nd, 3rd, 4th and beyond 4th week respectively.

**Figure 2.**
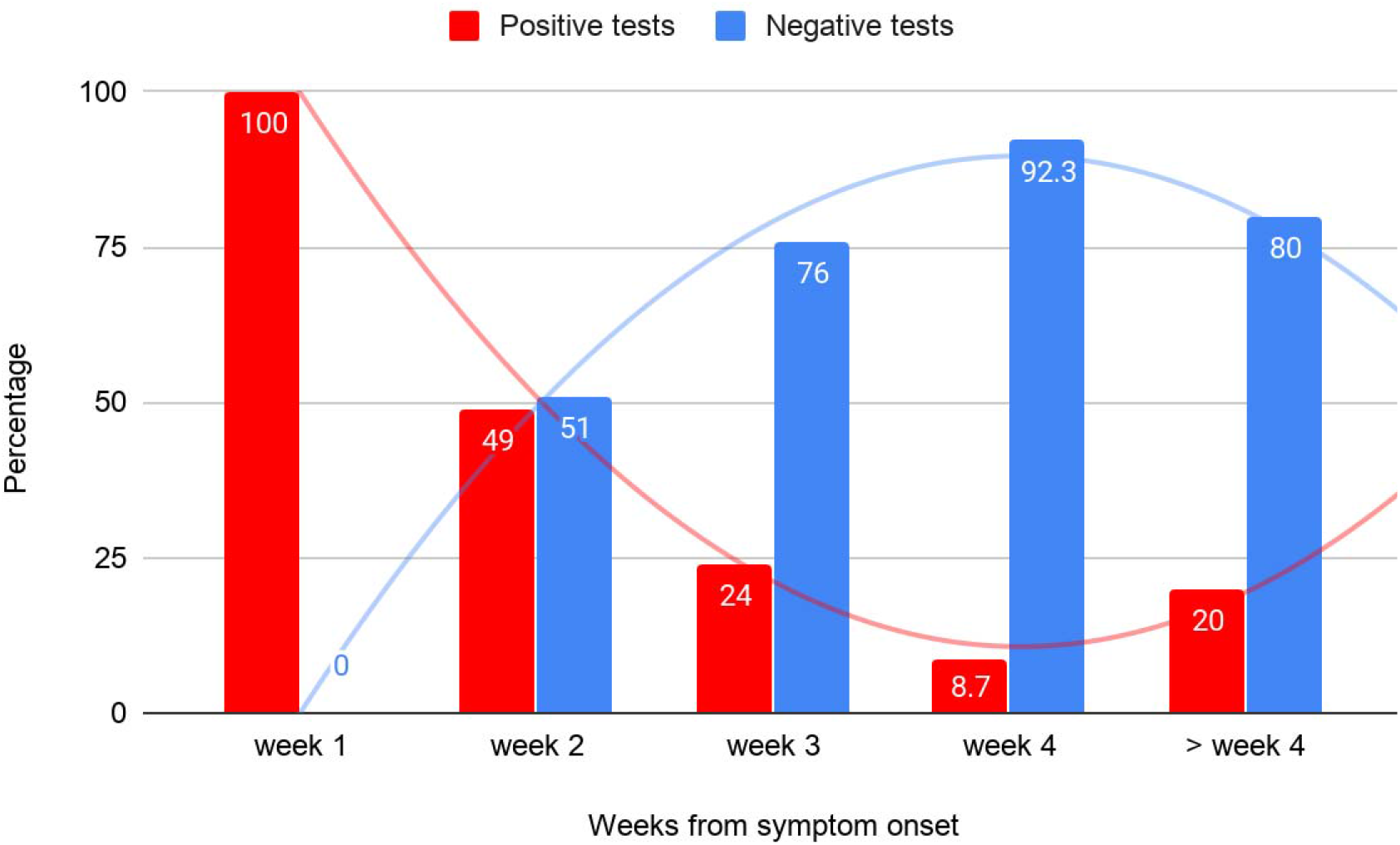
Percentage of positive and negative RT-PCR tests from day of symptom onset

Further analysis to look for association of age, gender, comorbidities, and symptomatology with the duration of RT-PCR positivity was done. The mean duration of positivity in age group 60 years or more, 19 - 59 years and <18 years were 22·33±7·17, 17·18±5·79 and 14·75±4·85 days (p= 0·04)(Figure 3). The age wise distribution of median duration has been given in Table 2. The mean period of last positive RT-PCR assay from the day of symptom onset among males and females (17·51±6·64 and 18·38±5·58 days, p=0·61), diabetics and non-diabetics (17·83±6·89 and 17·84±6·11 days,p=0·99), hypertensives and non-hypertensives (19·85±5·66 and 17·55±6·30 days, p=0.36). The mean duration from 1st and last positive RT-PCR assays between symptomatic and asymptomatic patients were 13·01±4·63 and 13·93±5·42 days respectively; and the difference was not statistically significant with p value <0·05 (p=0·39).

**Table 1.** Patient demographics (n=298)

| Variables | No. of patients (%) |
| --- | --- |
| Gender |  |
| Male | 226(75.83%) |
| Female | 72(24.10%) |
| Age (years) |  |
| 0-18 | 16(5.30%) |
| 19-59 | 254(85.23%) |
| ≥60 | 28(9.30%) |
| Presentation |  |
| Symptomatic | 164(55%) |
| Asymptomatic | 134(45%) |
| Comorbidities |  |
| Diabetes mellitus | 39(13.08%) |
| Hypertension | 23(7.70%) |

**Table 2:**
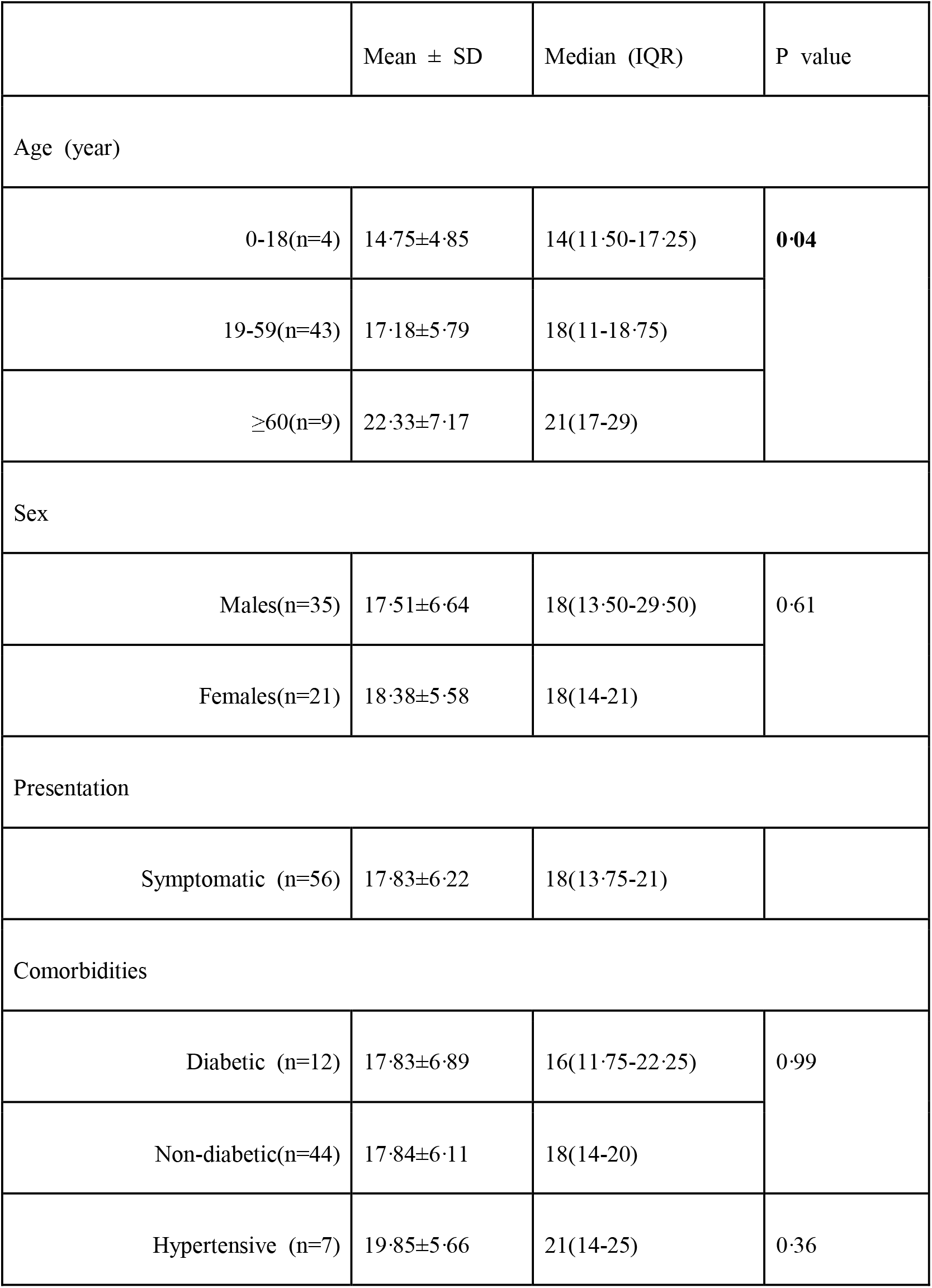
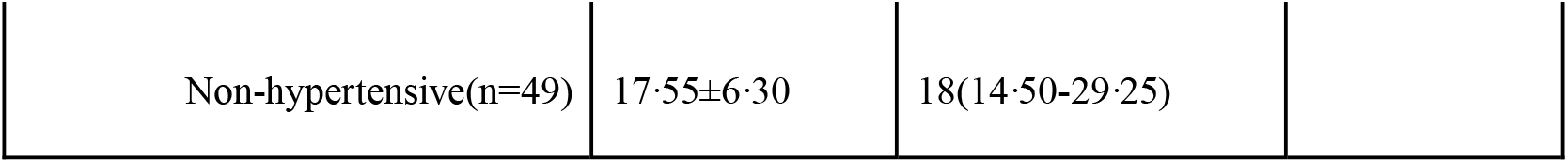
Duration of symptom onset to last positive RT-PCR (n=56)

**Figure 3.**
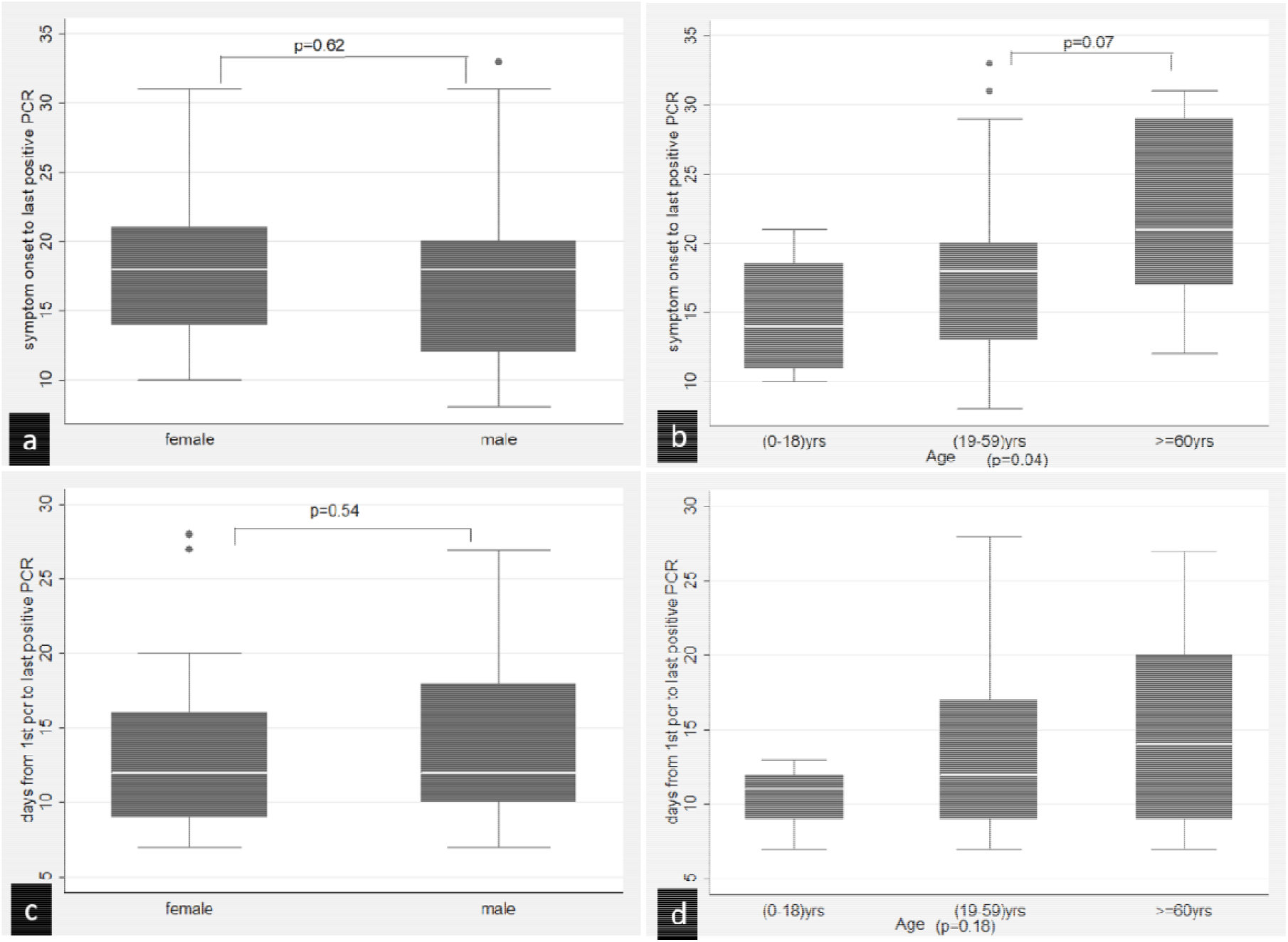
Box-plot analysis showing (A)Differences in duration of symptom onset to last RT-PCR positivity in (a) males and females (b) age groups 0-18 years, 19-59 years and ≥60 years (B) Differences in duration of first to last RT-PCR positivity (includes both symptomatic and asymptomatic case) in (c) males and females (d) age groups 0-18 years, 19-59 years and ≥60 years

In asymptomatic patients, mean duration of positivity (from 1st RT-PCR) among the 3 age groups,age 60 years or more, 19 - 59 years, < 18 years were 15(±6·59), 13·29 (±4·67), 10·5(±2·25) days [p value= 0·17]. The median time between 1st and last positive RT-PCR assays in the 3 age groups were 14(IQR:9-20), 12(IQR:9·25-17), and 11(IQR:9·25-12) days respectively (Table 3). There was no statistically significant difference among diabetics and non-diabetics (14·69±6·73 and 13·12±4·57 days, p=0·29), hypertensives and non-hypertensives (13·37±4·92 and 13·2±5·22 days, p=0·91), males and females (13·6±4·86 and 12·94±5·08 days, p=0·53).

**Table 3:** Duration of 1st to last positive RT-PCR (n=90)

|  | Mean(SD) | Median(IQR) | P value |
| --- | --- | --- | --- |
| Age (years) |  |  |  |
| 0-18(n=6) | 10.50±2.25 | 11(IQR:9.25-12) | 0.17 |
| 19-59(n=71) | 13.29±4.67 | 12(IQR:9.25-17) |  |
| ≥60(n=13) | 15±6.59 | 14(IQR:9-20) |  |
| Sex |  |  |  |
| Males(n=56) | 13.60±4.86 | 12(IQR:10-18) | 0.53 |
| Females(n=34) | 12.94±5.08 | 12(IQR:9-16) |  |
| Presentation |  |  |  |
| Asymptomatic(n=33) | 13.93±5.42 | 12(9-17) | 0.39 |
| Symptomatic (n=57) | 13.01±4.63 | 12(9-16) |  |
| Comorbidities |  |  |  |
| Diabetic (n=13) | 14.69±6.73 | 12(9-20) | 0.29 |
| Non-diabetic(n=77) | 13.12±4.57 | 12(9.25-18) |  |
| Hypertensive(n=10) | 13.37±4.92 | 10(9.25-16.50) | 0.91 |
| Non-hypertensive(n=80) | 13.20±5.22 | 12(10-18) |  |

Among symptomatic patients, 12 (7·3%) had positive RT-PCR beyond 21 days of symptom onset. Of these, 4 had diabetes mellitus, 3 had hypertension, and 4 were elderly i.e 60 years or above.

## Discussion

Understanding the pathogenesis, transmission dynamics of SARS CoV-2 is important to manage the cases, stop transmission and prevent further expansion of the pandemic. Prompt identification and isolation of cases is critical in order to halt the spread of infection. This study was conducted in the initial part of pandemic in India, which, to the best of our knowledge provides the largest data on SARS-CoV-2 RNA detection in naso-pharyngeal and oro-pharyngeal samples in symptomatic or asymptomatic COVID-19 cases.

As viral isolation is neither readily feasible nor available, diagnosis of viral RTIs relies upon molecular tests such as nucleic acid amplification and detection based tests. Moreover, the duration of viral shedding (or infectivity) in a clinical setting can only be ascertained by viral isolation in appropriate respiratory samples. However, in most cases viral nucleic acid detection and load (using cycle threshold, Ct value) may serve as surrogate marker for the same.

SARS-CoV-2 RT-PCR results have been the cornerstone of diagnosis as well as cure and discharge policies across different countries.(5) In addition RT-PCR assays have been utilised to determine the duration of persistence of the virus in respiratory tract samples. The median duration of viral detection in respiratory samples of mildly ill cases have been reported to be 14 days.(6) Also the viral loads in samples from severe cases were found to be higher than that of mild cases and 90% of mild patients cleared the virus, as reported by RT-PCR, within 10 days from symptom onset.(7) The exact period of infectivity of COVID-19 has been uncertain but transmission from asymptomatic patients has been documented.(8) In this cohort, patients were found to have detectable SARS-CoV-2 RNA via RT-PCR as early as 1 day after onset of symptoms with a median duration of 3 days(IQR:2-7) while median duration to last positive viral RNA detection was 18 days.

Further, few patients had prolonged detection of SARS-CoV-2 RNA i.e. longest duration being 33 days following symptom onset. Similar findings have also been noted by Xiao et al. in two published studies where they have highlighted the issue of prolonged viral nucleic acid detection.(9,10) Compared to viral isolation, RT-PCR tests have some intrinsic limitations. Firstly, COVID-19 infection cannot be ruled out based on negative RT-PCR results, particularly when highly suspected. False negative results can occur as a result of inadequate specimen or faulty sample collection technique, timing of sample collection, sample storage and processing and viral mutation.^(2)^ Further, detection of nucleic acid doesn’t imply presence of viable virus and therefore PCR positivity doesn’t necessarily relate to infectivity. A recent study by Scola et al. evaluated the correlation between Ct values of RT-PCR and viral isolation in cell culture where they found that patients with positive RT-PCR and Ct values more than 33-34 were not contagious.(11) Further studies on viability of the SARS-CoV-2 virus in respiratory samples are needed to draw a firm conclusion.

Association of age, sex, comorbidities, and symptomatology with duration of PCR positivity from 1st PCR or symptom onset was studied and analyzed. There was no statistically significant difference with respect to gender, presence of comorbidities (diabetes and hypertension). However, patients aged 60 years or more had a significantly prolonged duration of RT-PCR positivity since the onset of symptoms as compared to the younger age group. Xu et al. studied the patients with prolonged viral shedding in a retrospective study which revealed older age, male sex, corticosteroid treatment, delayed hospital admission and requirement of mechanical ventilation to be the associated risk factors. (12)The greater immune dysfunction in the older patients has been touted as a possible explanation for the prolonged persistence of SARS-CoV-2 in their respiratory tract^(10).^

A substantial number of cases in this study were asymptomatic patients. Identification of asymptomatic carriers is very important for public health measures considering the high basic Reproductive Number (R_0_->2·5) and high secondary attack rate of SARS-CoV-2 amongst household and close contacts.(13) Asymptomatic cases have gained special considerations due to persistence of virus in respiratory samples.(14,15) Due to lack of information regarding exact history of contact and time of exposure with a confirmed case, comparison of duration of RT-PCR positivity could not be made between asymptomatic and symptomatic patients. However, median duration between 1st and last positive RT-PCR was the same in both groups i.e. 12 days. Among asymptomatic patients, those aged 60 years or older had a longer median duration of positivity as compared to the other age groups though not statistically significant. Based on findings of overall increased positivity in older age groups, it is desirable to consider prolonged observation in elderly population; and repeat RT-PCR testing before declaring them completely cured. Though, the RT-PCR positivity was not significant statistically in cases with comorbidities, however such cases, being at high risk should be given similar consideration as elderly population.

This was a retrospective study where the RT-PCR results of naso-pharyngeal and oropharyngeal samples of COVID-19 patients were analyzed. The main strengths of this study were its large sample size and heterogeneous population. One of the limitations were inconsistent intervals of testing among patients, for which no definitive conclusion could be made about the actual nucleic acid conversion time. Due to lack of availability of viral isolation tests, the actual period of infectivity of patients couldn’t be determined, particularly those with prolonged positive results. More studies, comparing RT-PCR results with viral isolation are required to make unequivocal conclusions about viral shedding and infectivity.

## Conclusion

To the best of our knowledge this is the first study which provides information about the dynamic RT-PCR profile in COVID-19 cases in India. Most patients remain positive for SARS-CoV-2 after two weeks of symptom onset. Older age was associated with prolonged positivity. Patients with comorbidities and old age require special considerations from management and containment point of view.

## Data Availability

All data is secure and with the invetigators. It will be shared only when required.

## Author contribution

RK, BB, VM - conception of idea, study design, methods, data collection, data entry

VR, VM, MS - statistical analysis

BB, RK, VM - literature search, first draft of manuscript

VM, MS, NW - final draft, editing, proofreading

BB, RK, VM, MS, SV, NW, SB, AM - actively involved in management COVID-19 cases (part of institutional COVID task force)

VM - overall guarantor and corresponding author.

## Data availability

available and attached

